# Hematological Events in Metformin Treated NSCLC Patients Post Chemotherapy: Drug Repurposing Based on Real-World Evidence from Case Controlled Study

**DOI:** 10.1101/2025.04.26.25325377

**Authors:** Saikat Samadder

## Abstract

**Aim:** In this multi-center case controlled study aimed at metformin repurposing on the basis of survival benefit in non-small cell lung cancer patients compared to non-diabetic patients, based on hematological events mitigating ability of metformin induced by chemotherapy.

**Methods:** At the Severance hospitals from 1^st^ Jan 2010 to 1^st^ Jan 2020, (n = 1851) patients were identified from hospital database. Further analysis was performed using propensity score matched to analyze the survival outcome of metformin comparing with non-diabetic control group in non-small cell lung cancer patients. Using propensity score matching of 8 variables, post-chemotherapy days survived by diabetic patients compared with untreated patients shown in this study. Six different chemotherapy induced clinical events related to WBC, platelets, and neutrophils are studied to identify possible detrimental role of metformin post chemotherapy frequency/cycle stratification.

**Results:** Total 354/1851 patients identified as diabetic non-small cell lung cancer patients remaining were non-diabetic patients. Post-propensity score matching 354 non-diabetic patients survived 521 days (95% CI 459 – 582), compared to treatment group patients survived for 580 days (95% CI 508 – 651) non-significantly with (*p*-value 0.1). The patients under radiotherapy survived 624 days (95% CI 521 – 726) in 142 control group, compared to that chemoradiation metformin treated patients (n = 136) survived 798 days (95% CI 698 – 967) *p*-value-0.02. Among (n = 136) chemoradiation metformin patients treated prior to 180 days of chemotherapy survived mean days post chemotherapy start 2-year hazard ratio 0.42 (95% CI 0.008 – 0.7) *p-*value 0.02 compared to patients received metformin 180 days post chemotherapy. Post propensity score matching Metformin treatment did not significantly reduce leukopenia, neutropenia, and thrombocytopenia events post chemotherapy compared to control (*p*-value < 0.05). Further chemotherapy cycle stratification leukocytosis, neutrophilic leukocytosis, and thrombocytopenia events were significantly high (*p*-value < 0.05) causing increased deaths in chemotherapy non-responder compared to responder group.

**Conclusion:** The overall survival of diabetic patients treated with metformin is not different from untreated patients. Fatalities in treatment and control groups induced by chemotherapy were similar. Prolonged use of metformin reduced hematological event risk in NSCLC patients.

## INTRODUCTION

Lung Cancer is the second most cause of deaths among all cancers in the global population. As per GLOBOCAN report until 2020 a total of 1.7 million deaths due to lung cancer were recorded each year and 2.2 million newly diagnosed lung cancer were reported [1]. Several medicines were repurposed in past decades to counter this severe disease and to enhance the survival outcome [2, 3]. Anti-inflammatory medicine like celecoxib is presently prescribed for lung cancer patients originally known for the treatment of rheumatoid arthritis and ankylosing spondylitis. Pemetrexed is another best example of drug repurposed for lung cancer [4]. Maintaining the hematological balance within the clinical range was reported to have positive survival effect on cancer patients [5]. At the same time dual mode of actions such as immunosuppression and tumor growth inhibition is necessary to reduce adverse outcome in cancer. Identifying potential immunosuppressive agents similar to pemetrexed and celecoxib, beside immune-activating agents like GM-CSF, G-CSF, are the prime medications used to tackle chemotherapy induced immunopathologies [4, 6]. Events occurring from myelosuppression or hyperactivation may lead to detrimental conditions including death [7–12]. Chemotherapy induced diabetes is a rising concern and one of the longest ongoing debates to dissect the relationship between diabetes and cancer [12]. Type-II Diabetic patients were reportedly more prone toward various cancers depicting mainly gender, age, and lifestyle as the major confounding factors of disease progression [10].

Metformin, one of the oldest known medicines for diabetes, is known for increasing the risk of myelosuppression and related serious adverse events in long-term users [13]. In several retrospective analysis it was demonstrated that metformin use benefited cancer patient’s survival outcome [14, 15]. At the same time metformin was not found to reduce the risk of deaths in chemoradiation patients with advanced lung cancer [16, 17]. Myelosuppression and myeloupregualtion may occur due to ongoing or prolong chemotherapy regimen, or metformin use or due to cancer progression [18, 19, 20]. Therefore, events resulting in deaths of patients post chemotherapy end potentially after 1 to 2 years of standard first-line treatment, monitoring deceased patient’s immunopathological events retrospectively would potentially identify the actual cause of deaths in lung cancer population [20]. Events such as leukopenia, lymphocytopenia, neutropenia, leukocytosis, thrombocytopenia, and thrombocytosis are often caused due to cardiac events or infections, increases the overall disease burden [21, 22]. These events are often known as chemotherapy induced adverse events reduces quality of life. Diabetes is induced by cancer progression, chemotherapy, and radiation therapy as well [23].

The main focus of this propensity score matched (PSM) retrospective analysis is to measure and compare the hazard ratio of metformin users and non-users diagnosed with non-small cell lung cancer (NSCLC) for drug repurposing. In this study events and event-to-death occurrence in patients post chemotherapy initiation in metformin patients compared to non-diabetic patients is analyzed. Events such as leukopenia, lymphocytopenia, neutropenia, neutrophilic leukocytosis, thrombocytopenia, and thrombocytosis are compared in overall patients post-chemotherapy initiation to find the risk or benefit from metformin usage. Identifying different chemotherapy responders and finding the event related deaths is of great importance and are rarely performed for drug repurposing. Here it is further demonstrated using chemotherapy frequency/cycle stratified Kaplan-Meier plot the effect of chemotherapy responding NSCLC patient’s overall survival. By calculating the events occurrence rate and mortality rate in control and treatment groups as well in chemotherapy cycle stratified sub-groups, during 1^st^ year and 2^nd^ year post-chemotherapy initiation to identify underlying cause of fatalities in diabetic patients. As diabetic patients may endure maximum chemotherapy treatment response compared to non-diabetic cancer patients is presently not well-known necessary for drug repositioning against cancer.

## MATERIALS AND METHODS

### Study design and population

In this present multi-center case controlled study, the real world data (RWD) was obtained from three different branches of Severance hospital, consisting of electronic medical record (EMR) of patients receiving treatment for NSCLC. In this database patient information including age, gender, height, weight, body surface area (BSA), chemotherapy, immunotherapy, radiotherapy dose, dates of exposure, various clinical laboratory values obtained from continuous monitoring of hematological factors and patient vitals, various diseases and diagnosis dates, surgery and dates of surgical and radiological procedures are available, various non-chemo drugs, intake dates along with dose are accurately maintained. There is no missing information of patient included in this study.

To evaluate the effect of metformin in diabetic lung cancer patients aged above 19 years of age was included in this study. In hospital database there are 7,532 lung cancer patients identified based on searching this database with international classification of diseases-10 (ICD-10) codes among these patients (n=4,182) died until 30^th^ Jun 2024 received chemotherapy. Patients are excluded on following basis; missing chemo start date or chemo end date, SCLC and neuroendocrine lung cancer were excluded, missing WBC counts, neutrophil, RBC, platelet counts, small cell lung cancer patients, diagnosed with neuroendocrine tumor, and patients diagnosed with stage I/II during initial diagnosis were excluded. Among the deceased patients there are 2688 patients with available WBC counts were selected for analysis. By implementing further exclusion criteria, patients on chemotherapy treatment provided within 180 days post cancer diagnosis and patients receiving chemotherapy dose below 0.4 mg/m^2^. A total of (n=1851) patients were included in baseline, among these patients (n=354) were diagnosed with diabetes pre/post cancer diagnosis. The inclusion and exclusion criteria of patients in this study is shown using flowchart (Figure 1). Clinical baseline patients were selected based on the average count of 10 days pre/post cancer diagnosis allowing to include1851 patients. Post curation of data, the analysis was performed based on 1:5 PSM of 1851 patients including 354 samples in each arms allowing to obtain statistical strength of above 100%. The patients died during 1^st^ year and 2^nd^ year of chemotherapy start within 8-days post event occurrence were analyzed post-PSM and chemotherapy frequency/cycles stratification. This study design allowed to identify the cause of survival benefit or deaths arising from cancer progression, diabetes, chemotherapy, and/or metformin in diabetic and non-diabetic patients.

**Figure 1:**
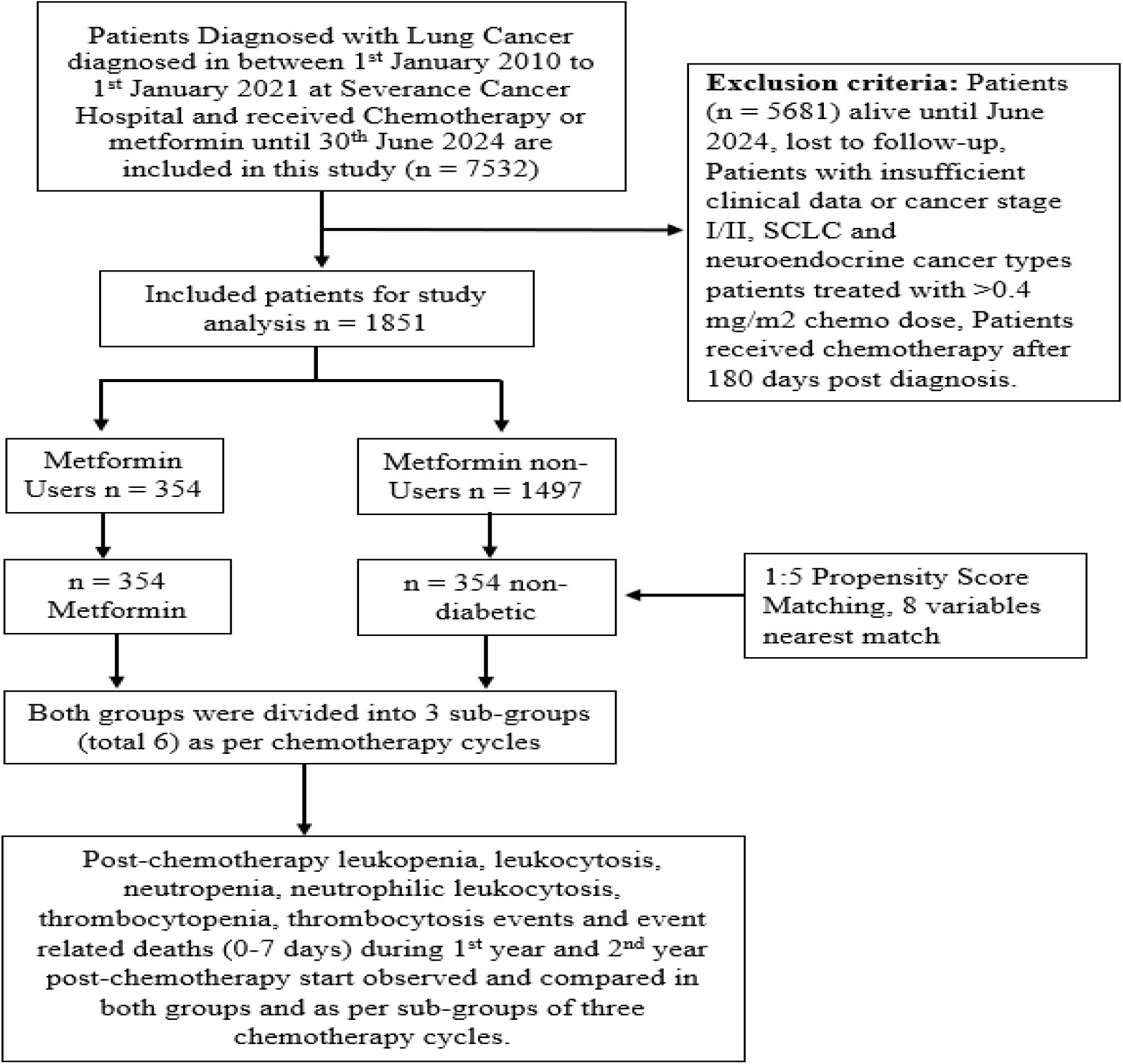
Study flowchart to identify myelosuppression and myeloupregualtion in Metformin treated/untreated Lung cancer patients. Three sub-groups were stratified based on chemotherapy frequency/cycle post propensity score matching to segregate chemotherapy response.

### Chemotherapy Dosage

To identify chemotherapy dose, volume of drug injected per cycle was divided from the BSA. The values of each patient’s BSA were pre-calculated and stored in database with high compliance for each patient. This allowed to acquire and calculate lung cancer patient total chemotherapy dosage as mg/m^2^ for all patients. Total dosage for oral formulation was calculated from the standard dose in mg available as per formulation type, multiplied by daily recommended dose, and duration of the treatment. Therefore, the total chemotherapy dosage was total of both mg and mg/m^2^ further utilized as a variable for PSM based on total chemo dosage [25]. This enabled to match patients with similar efficacy toward chemotherapy. While further stratification of chemotherapy frequency/cycle was performed to identify event occurrence and mortality rates among diabetic and non-diabetic groups.

### Propensity Score Matching

To perform PSM using 8 variables were included during the matching processes in RStudio. PSM was matched including gender, age, lung cancer location, stage, total chemo dosage, metformin treated/untreated, surgery, and radiotherapy status. The lung cancer metformin users ratio 1 were matched with non-diabetic metformin non-users with ratio 5; based on algorithm or values matched available from total chemotherapy dosage and 7 other variables [24, 25]. Obtaining standardized difference of between –0.1 to 0.1 post-PSM is considered as a sign of balance and was not found (supplementary figure **1-2**). PSM using chemotherapy dosage were rarely performed in previous literatures.

### Statistical Analysis

The statistical analysis was based on patients receiving all necessary therapies available for treatment post diagnosis as per patient’s requirement and physician’s decision. The covariates for this analysis are the age of cancer diagnosis, gender, body weight, cancer stage at diagnosis, lung cancer type, surgery, radiotherapy, salvage therapy, and total chemotherapy dosage. Responder and non-responders were analyzed by stratifying the frequency of chemotherapy post-PSM (shown in supplementary figure 5), in other terms it is the only possible method to study efficiency of chemotherapy. The overall survival curve of responders and non-responders would allow to understand the impact of metformin post-chemotherapy. Observing the change in hematological laboratory values could identify frequency of events and event-to-death within 1-year and 2-year post-chemotherapy start were analyzed in this study, provided with significant percentage change from baseline (shown in table **3** & **4**). Hazard ratio (HR) was calculated by dividing diabetic radiotherapy recipients died within 2 years with total number of patients, further dividing the ratio of patients died in Group A (180 pre chemotherapy start) by Group B (180 days pre/post chemotherapy start) and vise-versa for finding HR of Group A Vs Group C (180 days post chemotherapy start). Cox-proportional hazard regression analysis was used to estimate the HR of metformin users compared to non-users, with 95% confidence interval (CI). Data curation was performed in Microsoft excel, PSM and visualization were done with R (ver. 3.6.1; R Foundation for Statistical Computing, Vienna, Austria). The calculation of 95% confidence interval (95% CI), the *p*-value for mean and percentage was performed online using MedCalc [26].

**Table 1:**
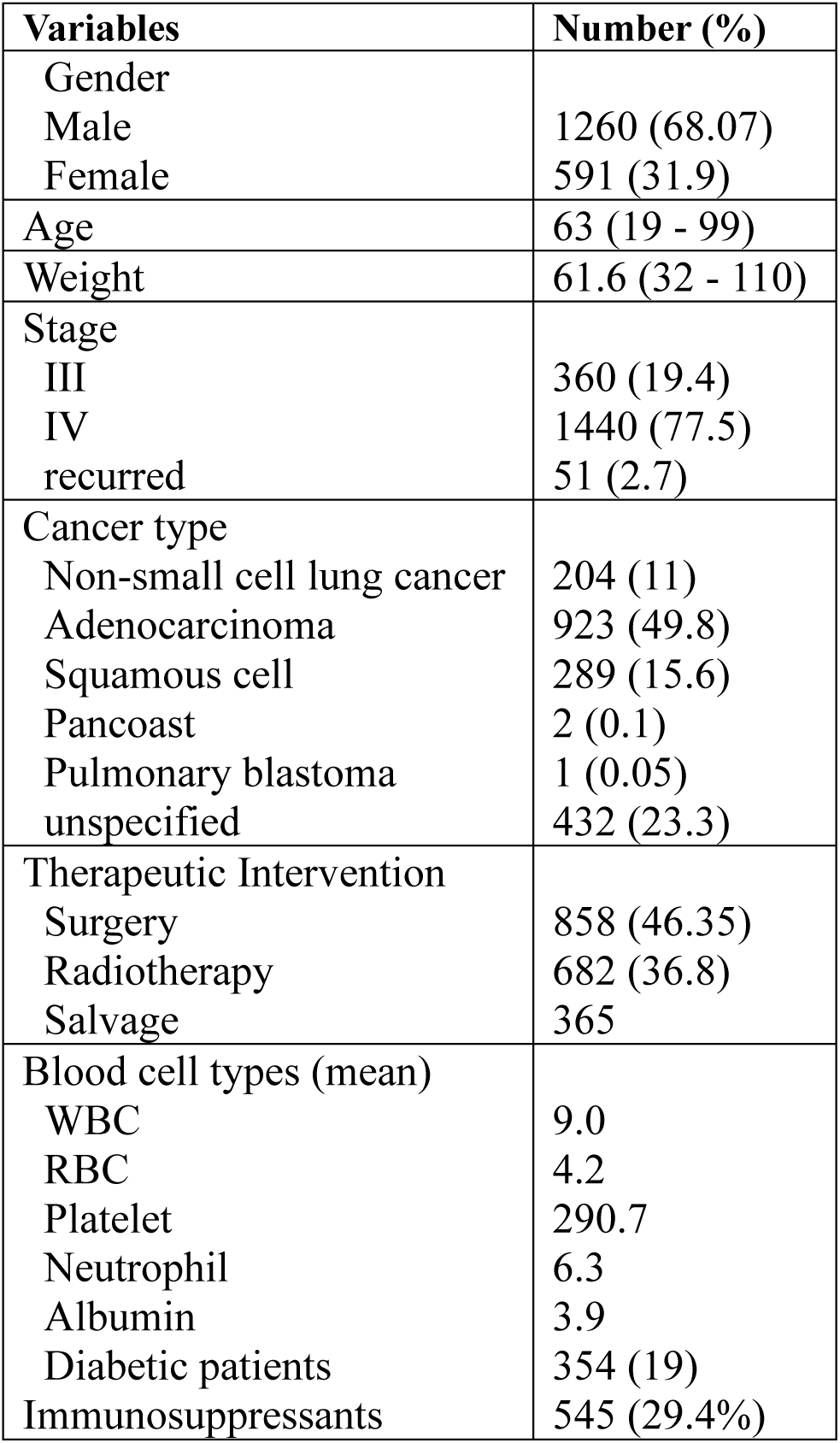
Pre-propensity score match baseline characteristics of NSCLC patients (n = 1851).

**Table 2:**
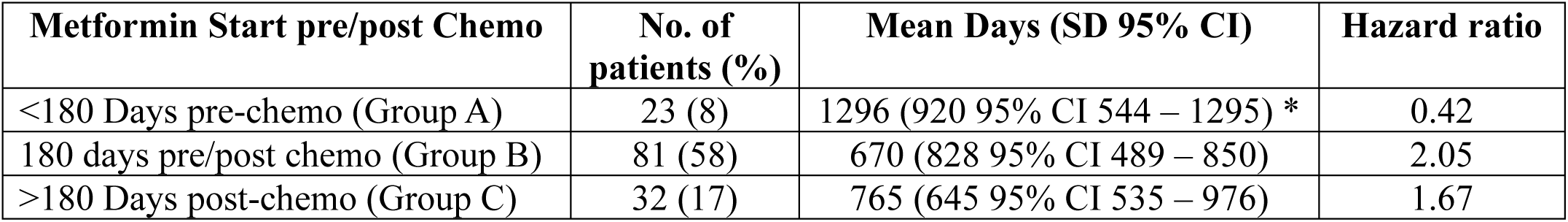
Sub-group survival analysis as per diabetes diagnosis and metformin treatment initiation in NSCLC chemoradiation patients. (*) represents statistical significance (*p*-value < 0.05) compared to Group B & Group C patients.

**Table 3:** Hematological events occurrence rate in NSCLC patients. Total events occurred at different timelines post cancer diagnosis, post chemotherapy start, compared between control and treated groups. (*) represents statistical significance (*p*-value < 0.05).

|  | Leukopenia | Leukocytosis | Neutropenia | Neutrophilic leukocytosis | Thrombocytopenia | Thrombocytosis |
| --- | --- | --- | --- | --- | --- | --- |
| **Total events pre/post chemotherapy, events (event %) (n = number of patients)** |  |  |  |  |  |  |
| **Control (n = 354)** | 2451 (46.25%)<br>(n = 264) | 3951 (50.2%)<br>(n = 310) | 1704 (46.45%)<br>(n = 244) | 5007 (48.2%)<br>(n = 334) | 4581 (54.7%)*<br>(n =260) | 1596 (47.5%)<br>(n = 202) |
| **Metformin (n = 354)** | 2848 (53.75%)*<br>(n = 267) | 3923 (49.8%)<br>(n = 313) | 1964 (53.5%)*<br>(n = 248) | 5373 (51.8%)*<br>(n = 337) | 3790 (45.3%)<br>(n =270) | 1764 (52.5%)*<br>(n = 206) |
| **Total events post chemotherapy initiation, events (event %) (n = number of patients)** |  |  |  |  |  |  |
| **Control (n = 354)** | 2168 (48.2%)<br>(n =252) | 3259 (52.3%)*<br>(n = 282) | 1469 (48.2%)<br>(n = 223) | 4092 (50.2%)<br>(n = 309) | 3358 (47.9%)<br>(n = 245) | 1227 (49.7%)<br>(n = 175) |
| **Metformin (n = 354)** | 2330 (51.8%)*<br>(n = 255) | 2972 (47.7%)<br>(n = 287) | 1581 (51.8%)*<br>(n = 229) | 4064 (49.8%)<br>(n = 320) | 3651 (52.1%)*<br>(n = 259) | 1239 (50.2%)<br>(n = 174) |
| **Post chemotherapy initiation 1<sup>st</sup> year, total events (event %) (n = number of patients)** |  |  |  |  |  |  |
| **Control (n = 354)** | 1456 (49.8%)<br>(n = 228) | 1899 (50.8%)<br>(n = 217) | 961 (51.3%)<br>(n = 205) | 2324 (49.7%)<br>(n = 250) | 2029 (48%)<br>(n =213) | 761 (47.8%)<br>(n = 143) |
| **Metformin (n = 354)** | 1468 (50.2%)<br>(n = 228) | 1833 (49.2%)<br>(n = 231) | 910 (48.7%)<br>(n = 206) | 2348 (50.2%)<br>(n = 273) | 2196 (52%)*<br>(n = 224) | 830 (52.2%)<br>(n = 140) |
| **Post chemotherapy initiation 2<sup>nd</sup> year, total events (event %) (n = number of patients)** |  |  |  |  |  |  |
| **Control (n = 354)** | 399 (53.9%)*<br>(n = 67) | 508 (56.7%)*<br>(n = 75) | 276 (52.5%)<br>(n = 58) | 743 (55.7%)*<br>(n = 93) | 590 (53%)*<br>(n =72) | 195 (45.4%)<br>(n = 41) |
| **Metformin (n = 354)** | 341 (46.1%)<br>(n = 64) | 389 (43.3%)<br>(n =76) | 249 (47.4%)<br>(n = 53) | 590 (44.3%)<br>(n = 90) | 522 (47%)<br>(n = 71) | 234 (54.6%)*<br>(n = 47) |

**Table 4:** 8-day standard mortality rate post event occurrence within 1^st^ year and 2^nd^ year post-chemotherapy initiation in (n = 708) NSCLC diabetic (metformin treated) Vs non-diabetic (untreated/control) patients. No statistical significance observed between the groups.

|  | Leukopenia | Leukocytosis | Neutropenia | Neutrophilic leukocytosis | Thrombocytopenia | Thrombocytosis |
| --- | --- | --- | --- | --- | --- | --- |
| **Total event-to-death pre/post chemotherapy, N = number of fatality (Mortality rate %)** |  |  |  |  |  |  |
| **Control (n = 354)** | N = 37<br>(10.4%) | N = 153<br>(43.2%) | N = 25<br>(7%) | N = 168<br>(47.4%) | N = 112<br>(31.6%) | N = 26<br>(7.3%) |
| **Metformin (n = 354)** | N = 29<br>(8.1%) | N = 143<br>(40.3%) | N = 21<br>(5.9%) | N = 173<br>(48.8%) | N = 119<br>(33.6%) | N = 23<br>(6.4%) |
| **Total event-to-death post chemotherapy initiation, n = number of fatality (Mortality rate %)** |  |  |  |  |  |  |
| **Control (n = 354)** | N = 37<br>(10.4%) | N = 153<br>(43.2%) | N = 25<br>(7%) | N = 168<br>(47.4%) | N = 112<br>(31.6%) | N = 26<br>(7.3%) |
| **Metformin (n = 354)** | N = 29<br>(8.1%) | N = 143<br>(40.3%) | N = 21<br>(5.9%) | N = 173<br>(48.8%) | N = 119<br>(33.6%) | N = 23<br>(6.4%) |
| **Post chemotherapy initiation 1<sup>st</sup> year event-to-death, N = number of fatality (Mortality rate %)** |  |  |  |  |  |  |
| **Control (n = 354)** | N = 23<br>(6.4%) | N = 96<br>(27%) | N = 15<br>(4.2%) | N = 103<br>(29%) | N = 65<br>(18%) | N = 14<br>(3.9%) |
| **Metformin (n = 354)** | N = 18<br>(5%) | N = 81<br>(22.8%) | N = 10<br>(2.8%) | N = 95<br>(26.8%) | N = 71<br>(20%) | N = 12<br>(3.3%) |
| **Post chemotherapy initiation 2<sup>nd</sup> year, total event-to-death, N = number of fatality (Mortality rate %)** |  |  |  |  |  |  |
| **Control (n = 354)** | N = 7<br>(2%) | N = 32<br>(9%) | N = 6<br>(2%) | N = 33<br>(9.3%) | N = 25<br>(7%) | N = 6<br>(2%) |
| **Metformin (n = 354)** | N = 4<br>(1%) | N = 20<br>(38%) | N = 2<br>(0.5%) | N = 29<br>(8.1%) | N = 17<br>(5%) | N = 6<br>(2%) |

### Event-to-Death Analysis

In the majority of patients event related mortality was observed within 0-7 days after 8 days, the mortality rate was high during that period. Therefore, the decision to analyze 0-7day mortality rate in patients was adopted. This fact was further confirmed in a separate analysis event-to-death per day reduced after 8 days (Day 0 to day 7) this confirmed post event 8 days was appropriate for analysis (supplementary figure 3). High mortality rate in the first month might not be specifically due to myelosuppression or other related events (supplementary figure 4). Standard event occurrence rate was calculated, dividing events in group by total events, event-to-death was calculated, dividing number of fatalities by total number of patients/groups. The normal laboratory range of WBC, platelets, neutrophil cell counts provided (supplementary table 2). Based on cell counts hematological events occurrence were considered as leukopenia (> 4×10^3^ cells/ul), lymphocytopenia (< 10×10^3^ cells/ul), neutropenia (> 2×10^3^ cells/ul), neutrophilic leukocytosis (< 7×10^3^ cells/ul), thrombocytopenia (> 150×10^3^ cells/ul), and thrombocytosis (< 400×10^3^ cells/ul).

## RESULTS

### Baseline Demographics

All deceased chemotherapy treated patients (n = 1851) with NSCLC identified from 1^st^ January 2010 to 1^st^ January 2020 were selected for baseline assessment. Patients died until 30^th^ June 2024 were considered as cut-off date prior to analysis (4.5 years). Among all 1851 were identified for analysis 68.07% male and 31.9% female, mean age at diagnosis was 63 years, mean body weight was 61.6 kg, all 360 (19.4%) patients at stage-III, 1440 (77.5%) were in stage-IV, remaining 51 (2.7%) were found recurred stage at baseline. In 1419/1851 (87.7%) patients were diagnosed with various types of non-small cell lung cancer during the first hospital visit remaining 432 (23.3%) were diagnosed as unspecified. Pulmonary surgical interventions were provided specifically for lung cancer (n = 858; 46.35%) patients, (n = 682; 36.8%) patients received radiation therapy, and (n = 365) patients were found to have undergone salvage therapy. At baseline in 1851 deceased individuals the WBC count was 9.0, RBC count was 4.2, platelet count was 290.7, neutrophil 6.3, albumin was 3.9 during the diagnosis of NSCLC. Post-PSM there were non-significant difference between the variables among groups (refer supplementary table 1) except albumin. Mean days of chemotherapy duration was 45 days less in control group compared to metformin treated patients identified post PSM.

Identifying and providing treatment for immunosuppressed patients prior to initiation of chemotherapy is highly recommended as immunosuppressant or chemotherapy may exacerbate myelosuppression mainly for diabetic patients. It was found that among 545/1851 patients and in matched 708 patients 230 patients received immunosuppressants at some point post diagnosis of cancer. Total 124/230 (53%) were in metformin group and 110/230 (47%) were treated with immunosuppressants in control group. Immunosuppressed 45/230 patients received immunosuppressants before 30 days of first-line chemotherapy that lasted for 6 months, 45 patients possibly experienced myeloupregualtion. Among 49/110 patients in control arm received immunosuppressants during similar timeline. Post chemotherapy initiation within 1-year 456 patients and 448 patients experienced leukopenia and leukocytosis, respectively. In 411 patients neutropenia, and 523 neutrophilia leukocytosis was distinctly observed. In 437 patients thrombocytopenia and 283 experienced thrombocytosis within 1-year post first-line therapy.

### Efficiency of Chemotherapy

Efficiency of chemotherapy was analyzed based on chemotherapy stratified dosage frequency. A small proportion of population died after the treatment start resulting in 380 deaths within 180 days post-chemotherapy initiation and these patients could be regarded as non-responder of chemotherapy. It was important to provide an overall survival curve (supplementary figure 5) to understand the efficiency of chemotherapy, based on stratification of patients as per response, as these patients completed at-least 3 to 6 months of first-line treatment and few presented better survival ability compared to non-responders, irrespective of chemotherapy dosage. Among 708/1851 matched patients responders survived maximum (n =58/708) 1351 days (95% CI 1160 – 1541) compared to non-responders (n = 380/708) 294 days (95% CI 254 – 333), and moderate-responders (n =270/708) survived 741 days (95% CI 658 – 823). PSM ensured partial randomization resulted in non-significant mortality difference observed between the groups of three sub-population caused by specific events (figure 5); results may vary in non-randomized cohort studies.

### Benefit from Metformin

Total 354/1851 patients identified as diabetic NSLC patients remaining are non-diabetic patients. Pre-PSM there was significant survival benefit in diabetic patients results not shown. Post PSM 354 non-diabetic patients survived 521 days (95% CI 459 – 582), compared to treatment group patients survived for 580 days (95% CI 508 – 651) with non-significant statistical difference *p*-value 0.1. Beyond 2-year survival post-chemo in control group matched patients in 77/354 patients survived mean 1392 days (95% CI 1228 – 1537) and in metformin group 85/354 patients survived 1580 days (95% CI 1418 – 1741) with *p*-value 0.1. Among 174/354 patients mean days survived 670 days (95% CI 569 – 770) received surgery in control group, compared to 176/354 patients in metformin 136 patients survived mean 745 days (95% CI 630 – 859) with non-significant *p-*value 0.1. The patients under radiotherapy survived 624 days (95% CI 521 – 726) post-chemotherapy (n = 142) control group patients, while in (n = 136) metformin treated patients survived 798 days (95% CI 698 – 967) with *p-*value-0.02. Among 136 diabetic patients diagnosed prior to 180 days of chemotherapy survived maximum days post chemotherapy start, 2-year HR 0.42 (95% CI 0.008 – 0.7) with *p-*0.02 compared to patients receiving chemotherapy and metformin post-chemo. The 2-year mean survival days is 322 in (n=106/142) untreated patients compared to 276 days (n=83/136) in metformin group, the 2-year hazard ratio of metformin Vs control group patient is 0.82 (95% CI 0.02 – 0.24) with *p-*value 0.01. Non-diabetic chemoradiation recipients survived 692 days (95% CI 576 – 806) post cancer diagnosis compared to diabetic chemoradiation patients survived 926 days (95% CI 768 – 1083) with statistical significance of *p-*value 0.009.

### Myelosuppression

Frequency of leukopenia, neutropenia, and thrombocytopenia was statistically significant in metformin patients (*p*-value < 0.05) compared to non-diabetic/control group patients post-chemotherapy, fatality was non-significantly low in metformin group. All these three events were chemotherapy induced events (figure 3 & 4). The risk of event occurrence rate increased significantly in metformin group for thrombocytopenia events after 1^st^ year of chemotherapy. There is no statistical significance in mortality rate observed between the control and metformin treated myelosuppressed patients within 1^st^ year or 2^nd^ year post chemotherapy start. During 2^nd^ year post-chemotherapy patients in control group experienced leukopenia and thrombocytopenia events rate increased significantly (*p*-value < 0.05) compared to metformin treated patients. The number of patients experienced events were not significantly different among the groups, the number of patients expired due to events were non-significantly lower in metformin group compared to control group (figure 4). Event-to-death prior to chemotherapy initiation did not occur in either of the groups for all hematological events.

### Myeloupregualtion

Total immune-hyperactivation events such as neutrophilic leukocytosis, and thrombocytosis was experienced significantly by metformin group patients (*p*-value < 0.05) pre/post initiation of chemotherapy. Myeloupregulation was non-significant in both diabetic and non-diabetic patients within 1^st^ year post-initiation of chemotherapy. In the 2^nd^ year thrombocytosis related events significantly increased in metformin treated patients (*p*-value < 0.05) compared to control. Leukocytosis and neutrophilic leukocytosis events increased significantly in control group patients (*p*-value < 0.05) than metformin treated patients. There is no statistical significance in mortality rate caused due to events observed between the control and metformin treated myeloupregulated patients. Although fatalities in metformin group was lower in leukocytosis, and neutrophilic leukocytosis events compared to treatment group except for thrombocytosis number of fatalities were not different among groups. The number of patients experiencing events, or event-to-death are not statistically significant among the groups (figure 3 & 4).

### Sub-group analysis

In this study 8-day mortality rate in 3 different sub-groups are shown in (**table 5**), sub-groups are divided as per efficiency toward chemotherapy cycles. Event-to-death occurrence rate, caused by leukocytosis, and thrombocytosis were absent in diabetic chemotherapy responder patients. No statistical significance observed in event-to-death between the sub-groups non-responders, moderate-responders, and responders between control Vs metformin groups. Diabetic patients not responding (n = 188) well towards chemotherapy were not at higher risk of death due to events related to myelosuppression and myeloupregulation compared to diabetic responders (n = 32). To confirm this event-to-death in all three chemotherapy groups of control and metformin are added (**table** 6) further divided by the total number of patients, as no statistical significance is observed in mortality rate between the control and treatment groups (figure 5). Total event-to-death in leukocytosis, neutrophilic leukocytosis, and thrombocytopenia were significantly high (*p*-value < 0.05) in non-responder compared to responder group (table 6).

**Table 5:** 8-day standard mortality rate post-chemotherapy in non-responder, moderate-responder and chemotherapy responders within 2 years of treatment initiation. Non-significant statistical difference observed between the three groups.

| Events | Non-responder (n = 380) |  | Moderate-responder (n = 270) |  | Responder (n = 58) |  |
| --- | --- | --- | --- | --- | --- | --- |
| (Total patients n = 708) | Metformin<br>(n=188) | Control<br>(n=192) | Metformin<br>(n=134) | Control<br>(n=136) | Metformin<br>(n=32) | Control<br>(n=26) |
| Leukopenia | 14 (7.4%) | 20 (10.4%) | 5 (3.7%) | 9 (6.6%) | 3 (9.3%) | 1 (3.8%) |
| Leukocytosis | 66 (35.1%) | 79 (41.1%) | 35 (26.1%) | 45 (33%) | 0 | 4 (15.3%) |
| Neutropenia | 7 (3.7%) | 13 (6.7%) | 3 (2.2%) | 7 (5.1%) | 2 (6.25%) | 1 (3.8%) |
| Neutrophilic Leukocytosis | 78 (41.48%) | 82 (42.7%) | 45 (33.5%) | 51 (37.5%) | 1 (3.1%) | 3 (11.5%) |
| Thrombocytopenia | 58 (30.8%) | 59 (30.7%) | 27 (20.1%) | 28 (20.5%) | 3 (9.3%) | 3 (11.5%) |
| Thrombocytosis | 12 (6.3%) | 10 (5.2%) | 6 (4.4%) | 9 (6.6%) | 0 | 1 (3.8%) |

**Table 6:**
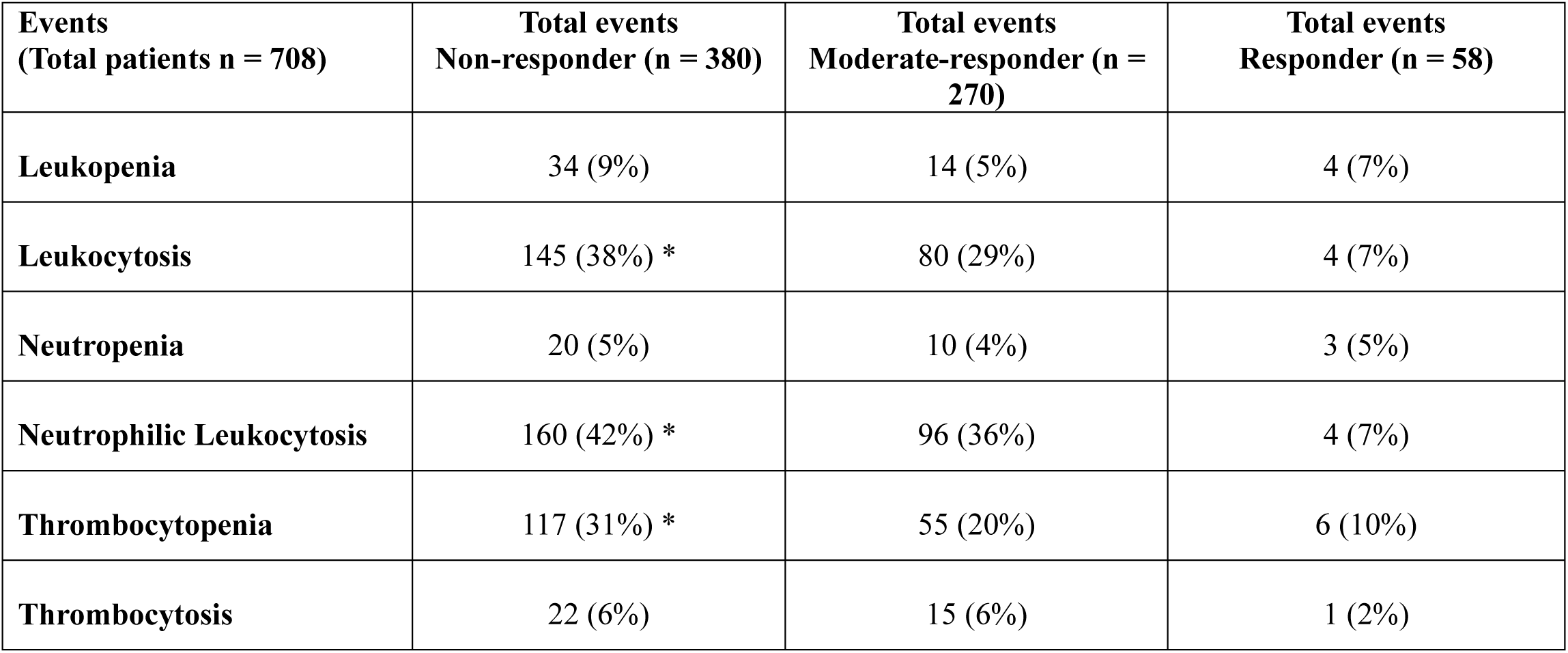
Total event-to-death in non-responder, moderate responder and responder sub-groups. (*) represents statistical significance (*p*-value < 0.05) comparison between non-responder Vs responder.

## DISCUSSION

In a recent randomized controlled trial (RCT), lung cancer patients showed no survival benefit from metformin and chemoradiation compared to patients treated with chemoradiation therapy [16]. In this study there is no overall survival benefit in diabetic NSCLC patients receiving metformin and chemotherapy compared to patients treated with chemotherapy. This study found diabetic chemoradiation recipients survived significantly longer compared to non-diabetic patients treated with chemoradiotherapy, because here most of diabetic patients received radiation therapy at the end stage and here survival was calculated post-chemotherapy start. Kim et al. reported maximum survival benefit in metformin treated squamous cell carcinoma patients, here adenocarcinoma patients are maximum [27]. It is confirmed in this study patients diagnosed with diabetes 6 months in prior to chemotherapy were less prone to fatality receiving chemotherapy, previously similar retrospective findings reported by Lee et al. in Asian diabetic NSCLC patients [28]. Patients responding well toward chemotherapy regimen were less prone to infections and bleeding caused by hematological events. These findings suggest that all responders blood cell sub-population were maintained within moderate range for prolong time post cancer diagnosis. Anti-tumor T-lymphocyte production and activation to eliminate tumor debris post-chemo, from the tumor site and in blood/lymph circulation possibly results in increased WBC activation [29]. Chemoresistance is one of the main causes of deaths in non-responder NSCLC patients, at the same time results in abnormal blood cell counts due to chemotherapy initiation are more likely related to cardiac events or infections [6, 19, 29]. In a prospective study recruiting cervical cancer patients found leukopenia as one of the leading causes of deaths post grade 2+ leukopenia events [7]. Prognosis of non-responders is one of the major challenges at present, identifying these patients as non-responders in prior to initiation of chemotherapy would benefit the overall survival of these patients. Several studies reported numerous biological markers, and cancer stage to be a good prognostic factor for survival, at present very few studies are available on prognosis of non-responder patients post cancer diagnosis prior to chemotherapy initiation [30]. Early detection of non-responders would allow selection of low dose metronomic chemotherapeutic regimen that would benefit patient’s treatment outcome. In this study NSCLC patients lived shorter duration post first-line chemotherapy in non-responders group, and these patients are usually withdrawn from chemotherapeutic regimen due to various adverse events further resulting in cancer progression.

In control group patients, the frequency of event occurrence was significantly low compared to diabetic patients, withdrawal of metformin for short duration might reduce the event occurrence rate, further study is necessary to confirm [27, 31]. Here it is found that a chemotherapy responder or non-responder could be a diabetic or non-diabetic patient. Several hematological events could be distinct in three different stratified chemotherapy frequency/cycle sub-population of NSCLC patients, no significant number of patients were found to be affected severely within stratified groups (table 5). To repurpose a drug the event-to-death caused by chemotherapeutic regimen initiated against cancer should be significantly low in treatment group compared to control, irrespective of chemotherapy response. At present there is no study available to confirm hematological marker assisted drug repurposing, mostly overall survival and disease free survival were major focus of all RCTs (32). Future studies should focus on hematological biomarkers for drug repositioning, mainly WBC counts. Prospectively designed clinical studies recruits non-diabetic cancer patients to study survival benefit from chemotherapeutic regimen concomitantly treated with metformin or other non-chemotherapeutic drugs for specific duration are considered for repositioning against cancer, at the same time effect on hematological markers due to prolong use of metformin in treatment-arm are not monitored (33). In prospectively designed RCTs, metformin failed to demonstrate survival benefit in lung cancer patients in the past decade (supplementary table 3). This study found that during the 2^nd^ year post chemotherapy the number of events reduced significantly in metformin group for most of the events except thrombocytopenia. Chemotherapy induced thrombocytosis is a biomarker for bleeding, or severe infections may lead to internal bleeding in patients (34-35). All the event-to-death were chemotherapy induced, and deaths were mainly caused due to infections and bleeding were significantly higher in non-responder group (table 6). Increased WBC and neutrophils counts indicates infection or body’s natural attempt to recover chemotherapy induced immunosuppression; neutropenia manifests sepsis, are related to chemotherapy use (36-37). Albumin levels were significantly higher in metformin group patients at baseline compared to control. It did not enhance efficacy of chemotherapy observed in events occurrence and mortality rate.

One of the main limitations of this study is one-to-many (1:M) PSM, this allowed to overcome the limitation of selection bias, and patients were not excluded from analysis post PSM. The types of chemotherapeutic agents are not provided in this study, beside that concomitant use of other drugs, and other comorbidities of patients were not included in the baseline or matched patients. In hospital, patients are generally treated with GM-CSF prior to the start or end of chemotherapy to reduce the risk of decreased WBC counts and related events that was not considered at baseline. In this study patient’s economic background, smoking, drinking, and health insurance status are important confounding factors were not taken into consideration. It is shown for the first time that post-PSM, segregating patients on the basis of chemotherapy cycles, it is possible to distinguish the safety concerns of secondary concomitant drug, arising from adverse events caused by either primary therapeutic agents (chemotherapy), or metformin for being a long-term prescribed drug. Decades ago, clinicians segregated patients as per chemotherapy response, later was found infeasible for adoption in trials due to lack of randomization (38). Therefore, survival benefit of metformin group compared to control is not shown here using Kaplan-Meier plot in responders, moderate responders, and non-responders sub-groups. Moreover, there is no difference in survival between control and treated patients in these stratified sub-populations. All together metformin use is safe for diabetic NSCLC patients and the event related to death were not associated with metformin use. During the 1^st^ year post chemotherapy number of events increased but did not majorly contribute to deaths of diabetic patients. Further study is required to understand the reason behind this trend in metformin treated diabetic patients not found in non-diabetic control patients. In previous studies, mortality was not related to neutropenia or thrombocytopenia in cancer patients treated with metformin [35, 39–40]. In contrast this study found total neutropenic events post chemotherapy were significantly higher than control, but metformin did not contribute to high mortality rate. Total leukopenia, neutropenia, and thrombocytopenia events total occurrence rate were significantly higher post chemotherapy but significantly reduced in the 2^nd^ year, possibly due to prolonged use of metformin possibly reduced risk of hematological events. This fact could be further confirmed in overall survival benefit of NSCLC patients diagnosed with diabetes (< 180) days prior to chemotherapy initiation. Diabetic patients are often at risk of low platelet count due to high glucose level in blood. In overall diabetic patients metformin could not be considered as a major causative agent of thrombocytopenia or associated deaths in NSCLC patients. Here hematological events associated with chemotherapy or cancer progression resulted in event related deaths of NSCLC patients. Metformin attributed to increased event occurrence rate did not cause increased deaths caused by these events, suggesting metformin use is safe, withdrawal of metformin for short duration post severe event occurrence during ongoing chemotherapeutic regimen may increase therapeutic efficacy. The efficacy of metformin in survival outcome of NSCLC patients is beneficial for patients diagnosed with diabetes and treated prior to chemotherapy initiation. Metformin could be beneficial in other cancer types comorbid with diabetes, significant survival benefit could not be observed in NSCLC patients for metformin to be repositioned against NSCLC.

## Supporting information

STROBE

## Acknowledgement

The author would like to thank professor Kyungsoo park (Department of pharmacology, Yonsei University) for providing access to severance hospital database for obtaining patient’s electronic health records.

## Ethics Statement

I confirm all relevant ethical guidelines have been followed, and any necessary IRB and/or ethics committee approval was obtained. The studies involving human participants were waived by the Institutional review board (IRB) of Yonsei University. Written informed consent form for participation was not required for this study in accordance with the national legislation and the institutional requirements.

## Data availability

Data will be made available upon request, post-approval from Yonsei University IRB.

