## Supplementary for "Hematological Events in Metformin Treated NSCLC Patients Post Chemotherapy: Drug Repurposing Based on Real-World Evidence from Case Controlled Study"

### Hematological Events in Metformin Treated NSCLC Patients Post Chemotherapy: Drug Repurposing Based on Real-World Evidence from Propensity Score Matched Analysis

Saikat Samadder[1*]

[1*]Corresponding author; Department of Pharmacology, Yonsei University College of Medicine, 50 Yonsei-ro, Seodaemun-Gu, Seoul, Korea.


#### Supplementary Material

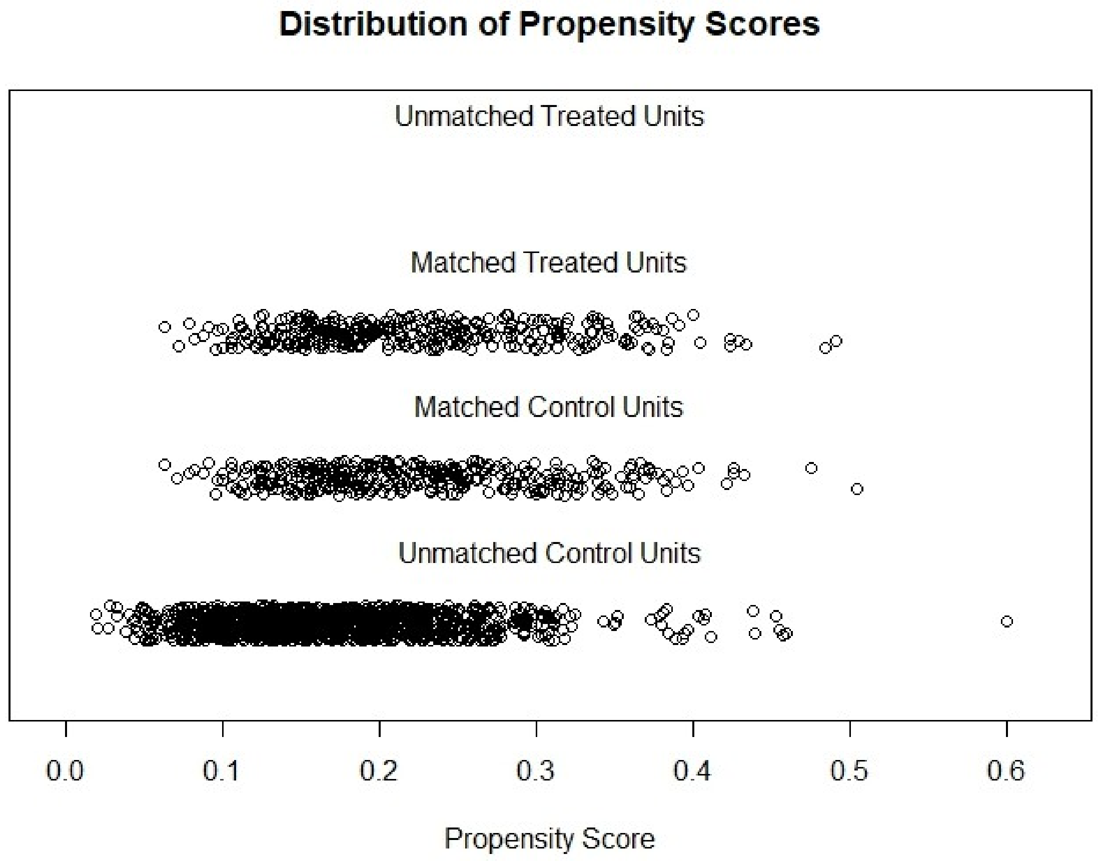


**Supplementary figure 1:** Distribution of 1:5 propensity score matching in metformin treated Vs control group patients.

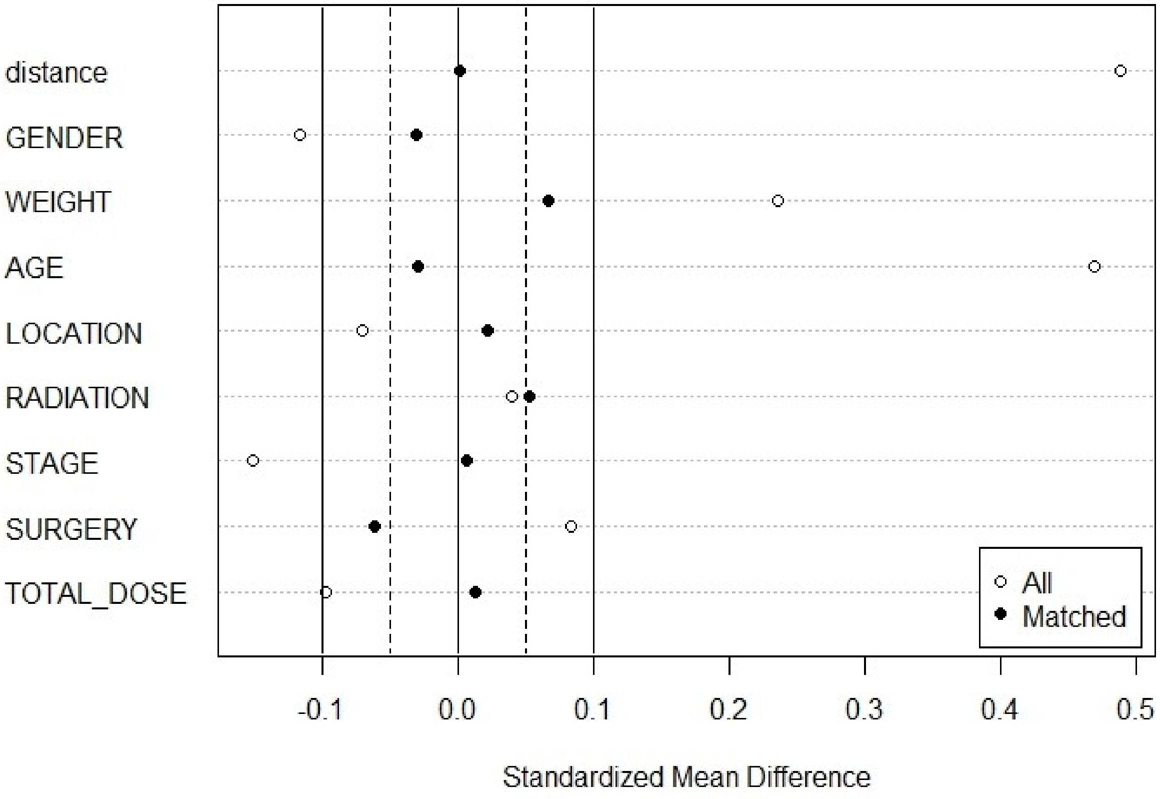


**Supplementary figure 2:** Standardized mean difference in 8 variables used in propensity score matching.

| | **Metformin (n = 354)** | **Control (n = 354)** | ***p*-value** |
|---|---|---|---|
| **Age** | **66.8 (SD 8.65)** | **66.9 (SD 10.21)** | **-** |
| **Gender** | | | |
| Male | **256 (72%)** | **259 (73%)** | **-** |
| Female | **98 (28%)** | **95 (27%)** | **-** |
| **Weight** | **63.5** | **63.6** | |
| **Cancer Stage** | | | |
| III | **91 (26%)** | **93 (26%)** | **-** |
| IV | **256 (72%)** | **254 (72%)** | **-** |
| recurred | **7 (2%)** | **7 (2%)** | **-** |
| **Cancer type** | | | |
| Non-small cell lung cancer | | | |
| Adenocarcinoma | **232 (66%)** | **245 (69%)** | **-** |
| Squamous cell | **80 (23%)** | **64 (18%)** | **-** |
| unspecified | **42 (11%)** | **45 (13%)** | **-** |
| Therapeutic Intervention | | | |
| Surgery | **176 (50%)** | **174 (49%)** | **-** |
| Radiotherapy | **136 (38%)** | **142 (40%)** | **-** |
| Salvage | **76 (22%)** | **76 (22%)** | |
| Blood cell types (mean) | | | |
| WBC | **8.9 (SD ± 2.93)** | **8.8 (SD ± 3.8)** | **-** |
| RBC | **4.2 (SD ± 0.52)** | **4.2 (SD ± 0.53)** | **-** |
| Platelet | **283.9 (SD ± 93.5)** | **286.5 (SD ± 95.8)** | **-** |
| Neutrophil | **6.1 (SD ± 2.7)** | **6.2 (SD ± 3.4)** | **-** |
| Albumin | **3.9* (SD ± 0.54)** | **3.8 (SD ± 0.53)** | **< 0.05** |
| Immunosuppressants | **100 (28%)** | **111 (31%)** | **-** |

**Supplementary table 1:** Post propensity score match baseline characteristics of (n = 708) patients. SD - stands for standard deviation. (*) represents statistical significance ($p$-value < 0.05)

| | WBC | Neutrophil | Platelet |
|---|---|---|---|
| Lower range | > 4 x $10^3$ cells/ul | > 2 x $10^3$ cells/ul | > 150 x $10^3$cells/ul |
| Normal range | 4 – 10 x $10^3$ cells/ul | 2 – 7 x $10^3$ cells/ul | 150 – 400 x $10^3$cells/ul |
| Higher range | < 10 x $10^3$cells/ul | < 7 x $10^3$ cells/ul | < 400 x $10^3$cells/ul |

**Supplementary table 2:** Normal, high and low ranges of WBC, neutrophil and platelet counts.

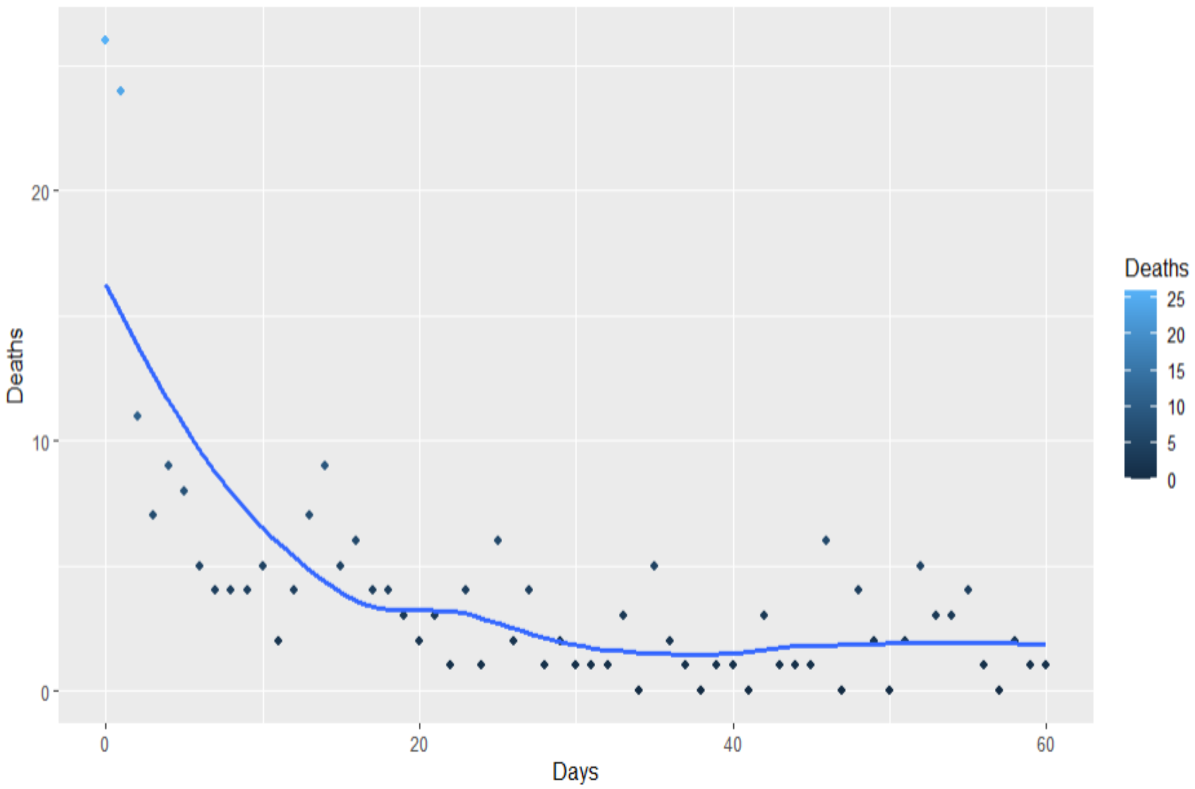


**Supplementary figure 3:** Total number of deaths after chemotherapy initiation leukopenia after two months in NSCLC patients.

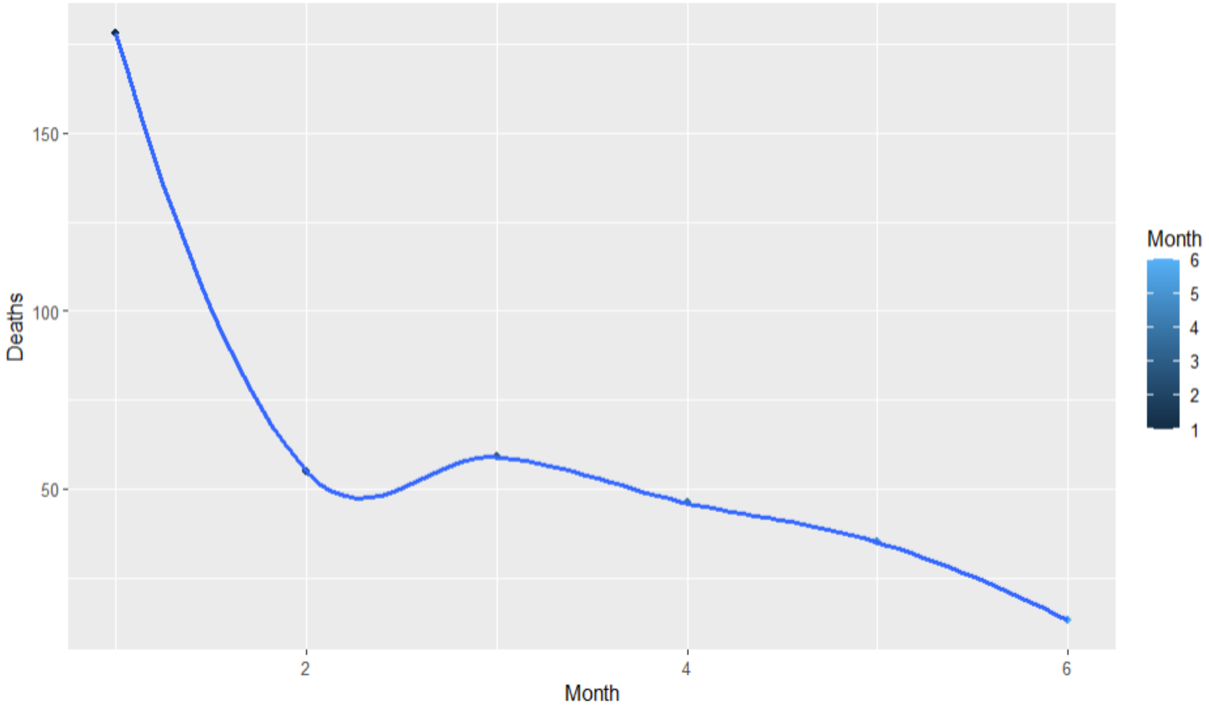

**Supplementary figure 4:** Decline in number of deaths post leukopenia in Lung cancer patients.

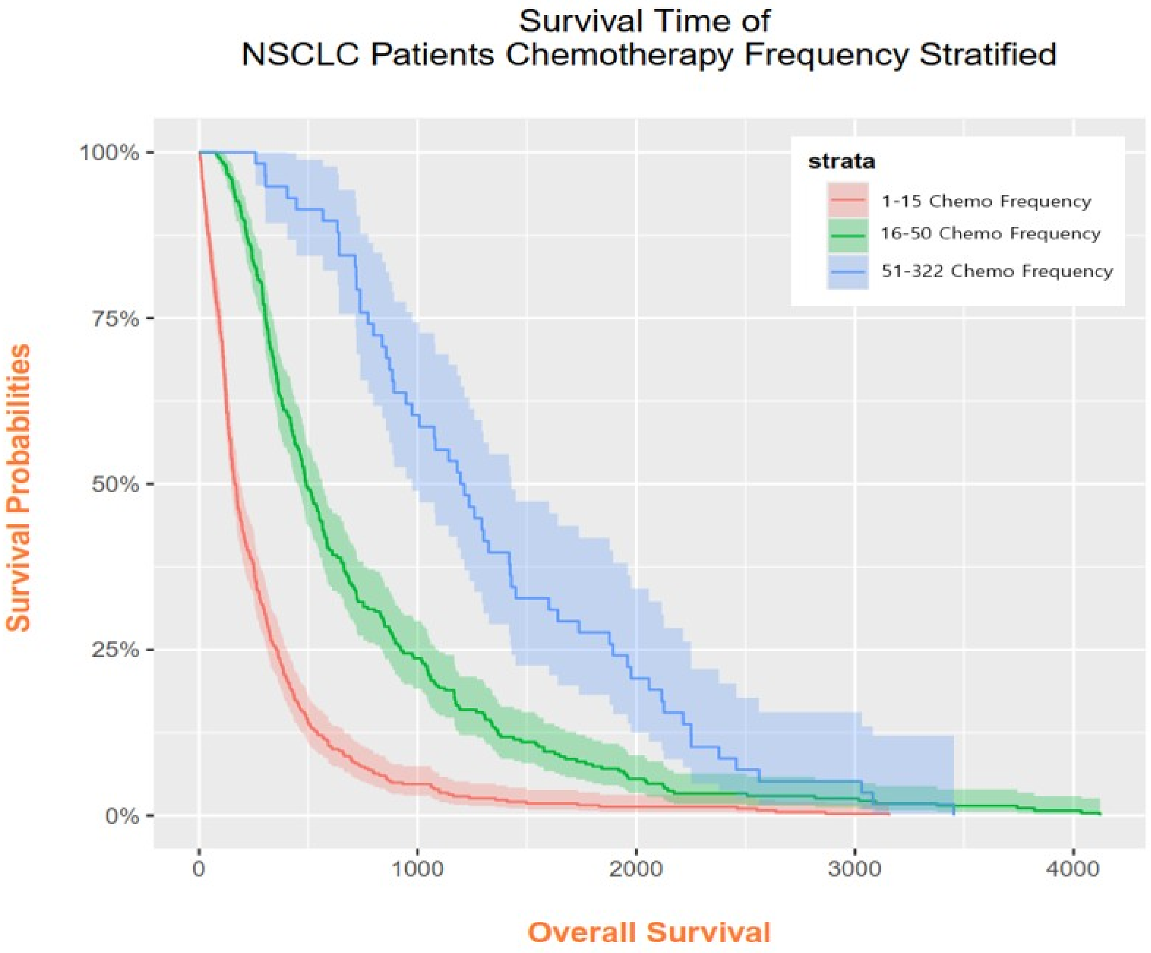


**Supplementary figure 5:** Chemotherapy frequency stratified survival analysis in (n = 708) NSCLC patients.

| Drugs | Indication/ Class | Approval year by FDA | Cancer type | Reference from literature review | Initial Approach to reach clinical studies | Current phase in Clinical trial | Approval status for cancer | Country of Origin | Published Results available |
|---|---|---|---|---|---|---|---|---|---|
| Metformin | Type 2 Diabetes mellitus | 1972 | Lung cancer | **1-29** | Preclinical, Retrospective analysis | NCT02115464,<br>NCT02285855,<br>NCT02019979,<br>NCT01717482,<br>NCT01997775,<br>NCT04170959,<br>NCT01578551,<br>NCT03086733,<br>NCT02109549<br>NCT01864681<br>NCT00659568<br>NCT02145559<br>NCT02431676 | Phase-II terminated<br>Phase-II terminated<br>Phase-II terminated<br>Phase-II terminated<br>Phase-II terminated<br>Phase-II terminated<br>Phase-II terminated<br>Phase-II completed<br>Phase-II completed<br>Phase-II completed<br>Phase-I completed<br>Phase-I completed<br>Phase-II completed | Canada<br>USA Latino 88%<br>Israel<br>USA Latino 100%<br>Taiwan<br>Belgium<br>USA<br>Canada<br>Netherlands<br>China<br>UK<br>USA<br>USA Black 45% | **30-37** |

**Literature search results of randomized controlled trials for lung cancer treated with Metformin and chemotherapeutics.**

**Supplementary Table 3:** Clinical trials status various cancers treated with chemotherapy and Metformin. Here trials with status of completed, unknown and terminated are presented.
